# Circulating tumor DNA analysis in advanced urothelial carcinoma: insights from biological analysis and extended clinical follow-up

**DOI:** 10.1101/2023.06.20.23291650

**Authors:** Sia Viborg Lindskrog, Karin Birkenkamp-Demtröder, Iver Nordentoft, George Laliotis, Philippe Lamy, Emil Christensen, Derrick Renner, Tine Ginnerup Andreasen, Naja Lange, Shruti Sharma, Adam ElNaggar, Minetta C. Liu, Himanshi Sethi, Alexey Aleshin, Mads Agerbæk, Jørgen Bjerggaard Jensen, Lars Dyrskjøt

**Author notes:** Corresponding author: Prof. Lars Dyrskjøt, Department of Molecular Medicine, Aarhus University Hospital, Palle Juul-Jensens Boulevard 99, 8200 Aarhus N, Denmark.

## Abstract

**Purpose:** To investigate whether circulating-tumor DNA (ctDNA) assessment in patients with muscle-invasive bladder cancer predicts treatment response and provides early detection of metastatic disease.

**Experimental Design:** We present full follow-up results (median follow-up: 68 months) from a previously described cohort of 68 neoadjuvant chemotherapy (NAC)-treated patients who underwent longitudinal ctDNA testing (712 plasma samples). In addition, we performed ctDNA evaluation of 153 plasma samples collected before and after radical cystectomy (RC) in a separate cohort of 102 NAC-naïve patients (median follow-up: 72 months). Total RNA-sequencing of tumors was performed to investigate biological characteristics of ctDNA shedding tumors.

**Results:** Assessment of ctDNA after RC identified metastatic relapse with a sensitivity of 94% and specificity of 98% using the expanded follow-up data for the NAC-treated patients. ctDNA dynamics during NAC was independently associated with patient outcomes when adjusted for pathological downstaging (HR=4.7, *p*=0.029). For the NAC-naïve patients, ctDNA was a prognostic predictor before (HR=3.4, *p*=0.0005) and after RC (HR=17.8, *p*=0.0002). No statistically significant difference in recurrence-free survival for patients without detectable ctDNA at diagnosis was observed between the cohorts. Baseline ctDNA positivity was associated with the Ba/Sq subtype and enrichment of epithelial-to-mesenchymal transition and cell-cycle associated gene sets.

**Conclusions:** ctDNA is prognostic in NAC-treated and NAC-naïve patients with more than five years follow-up and outperforms pathological downstaging in predicting treatment efficacy. Patients without detectable ctDNA at diagnosis may benefit significantly less from NAC, but additional studies are needed.

## Introduction

Bladder cancer is a common malignancy with more than 570,000 new cases each year worldwide (1), of which approximately 25% are diagnosed with muscle-invasive bladder cancer (MIBC). Despite curative intent treatment with neoadjuvant chemotherapy (NAC) followed by radical cystectomy (RC), approximately 50% of patients with localized MIBC develop metastatic disease (2) and only 40-45% have a pathological response to NAC (3,4). Early detection of metastatic relapse and effective monitoring of treatment response are therefore critical to improve patient outcomes.

Circulating tumor DNA (ctDNA) is a promising minimally-invasive blood-based biomarker for early detection of metastatic relapse and monitoring of treatment response in bladder cancer (5–7). Our group has previously demonstrated that ctDNA monitoring in patients with MIBC (median follow-up of 21 months after RC) can identify metastatic relapse with a median lead-time of 96 days over radiographic imaging (5). Several factors, including tumor stage and tumor size impact ctDNA detection (8–11). However, absence of ctDNA detection in advanced tumors of high volume has shown to be influenced by biological features such as age, obesity and diabetes (12) and also tumor characteristics including histology and proliferation rates (13). Thus, establishment and validation of sensitive ctDNA detection methods and increased knowledge of tumor characteristics affecting ctDNA shedding in bladder cancer are needed to facilitate the implementation of ctDNA assessment in routine clinical practice. As late events may be more difficult to detect due to continued metastatic tumor evolution (14), evaluation of ctDNA-based stratification of patients having long-term clinical follow-up is of high importance. Here we present extended clinical follow-up results (median follow-up of 68 months after RC) from a previously described cohort of 68 NAC-treated patients (5). In addition, we performed ctDNA evaluation in a retrospectively collected cohort of 102 patients that did not receive NAC to compare recurrence rates between NAC-treated and NAC-naïve patients when stratified by ctDNA status. We investigated whether longitudinal ctDNA assessment in patients with MIBC predicts treatment response and early detection of metastatic relapse for both cohorts. Furthermore, we evaluated the underlying biology of metastatic relapse and ctDNA shedding using RNA-sequencing (RNA-seq) data of the primary tumors at diagnosis.

## Materials and Methods

### Patients and clinical samples

All patients provided written informed consent, and the study was approved by The National Committee on Health Research Ethics (#1302183 and #1706291). Details of the 68 NAC-treated patient cohort have previously been described (5). Pathological downstaging after NAC was defined as ≤pTa,CIS,N0. Additional ctDNA analysis of 56 plasma samples collected after RC was performed.

For the NAC-naïve cohort, we retrospectively included 102 patients diagnosed with MIBC who underwent RC without prior treatment with NAC at Aarhus University Hospital in Denmark between 2001 and 2014. Patients were recruited over a period of 13 years (2001–2014) when NAC was not standard-of-care in Denmark. Hence, the cohort might include both cisplatin-eligible and ineligible patients. The NAC-naïve cohort was selected to represent an equal number of patients with and without metastatic disease within the cohort, and was not matched to the NAC-treated cohort. Plasma samples collected before RC (at diagnosis or at previous visits due to NMIBC) and after RC were included and analyzed for presence of ctDNA (collection of samples between 2001 and 2016).

Clinical end points were obtained from radiographic imaging results and pathology reports (recurrence-free survival [RFS]) and from the nationwide civil registry (overall survival [OS]). RC was not completed for patients 4175 and 4250 and recurrence assessment was not available for patient 4519, and these patients were therefore excluded from analyses of recurrence status. For patients in the NAC-naïve cohort, RFS was censored after 4 years of FU. Assessment of 12-month recurrence status was based on imaging data up to 14 months after RC to allow for variability in scheduling of imaging. Follow-up information was collected and managed using REDCap hosted at Aarhus University (15,16).

### Whole-exome sequencing

Whole-exome sequencing (WES) was performed on tumor embedded in Tissue-Tek® O.C.T.™ Compound or formalin-fixed, paraffin-embedded (FFPE) tumor tissue along with matched PBMC blood samples (germline) from each patient. Details and metrics of the NAC-treated 68-patient cohort were previously described(5). For the NAC-naïve cohort, WES of tumor and matched germline DNA was performed at a mean target coverage of 405X (range: 238-689X) for tumor samples and 94X (range: 60-139X) for germline samples.

### Personalized ctDNA assay using multiplex PCR (mPCR)-based NGS workflow

Personalized, tumor-informed ctDNA assays (Signatera^TM^) were designed as previously described (5) and used for ctDNA detection and quantification. Briefly, up to 16 high-ranked, patient-specific, somatic, single nucleotide variants (SNVs) derived from WES of tumor tissue were selected for multiplex (m)PCR testing. mPCR primers targeting the selected SNVs were designed, synthesized, and used for longitudinal ctDNA assessment. Plasma samples with at least two variants detected were defined as ctDNA-positive. ctDNA concentration was reported as mean tumor molecules (MTM) per mL of plasma.

Details and metrics of the NAC-treated 68-patient cohort were previously described (5). For the NAC-naïve cohort, a median of 3.6 mL of plasma (range: 1.3-7.9 mL) was used for cfDNA extraction. A median of 6.5 ng cfDNA per mL plasma (range: 0.79-799.8 ng/mL) was extracted. Baseline ctDNA status was defined as the ctDNA status pre-NAC for the NAC-treated cohort and the ctDNA status pre-RC for the NAC-naïve cohort. Accumulated ctDNA status after RC was defined as any ctDNA positive plasma sample during the surveillance period after RC. In addition, the accumulated ctDNA status within one year after RC was included to illustrate a defined time frame of evaluating patient prognosis after RC.

### Genomic characterization of tumors

The WES data of tumors from both patient cohorts were subsequently and separately analyzed for tumor characterization purposes (unrelated to the Signatera WES and personalized SNV assay design workflow). Reads were mapped with bwa-mem using the GRCh38 genome assembly, and SNVs and indels were called using Mutect2 (v2.2) and annotated using SnpEff (v4.3t). Variants with more than 2 alternate allele reads in the germline, less than three alternate allele reads in the tumor or a tumor VAF below 5% were filtered out. All variants passing the above filters were included in the analysis of mutational signatures using SomaticSignatures (v2.30) and MutationalPatterns (v3.4.1). Tumors with <50 SNVs were excluded (23/170 tumors). Trinucleotide patterns for COSMIC signatures (v3.2) were obtained and used for analysis of the contribution of the four main signatures previously observed in bladder cancer (SBS1, SBS2, SBS5, SBS13), rather than performing *de novo* extraction of signatures. To ensure that the contribution of the selected COSMIC SBS signatures was representative of the observed mutational spectrum, the resulting trinucleotide mutational profile for every sample was compared to the original profile and only samples with a cosine similarity above 0.9 were considered (107/147 tumors). For the analysis of the number of SNVs and indels according to baseline ctDNA status, only variants with a high or moderate impact (based on SnpEff annotation) were included. When assessing the mutation rates of single genes between ctDNA positive and -negative patients, a curated list of 78 bladder cancer driver genes from IntOGen was used.

### Total RNA-sequencing and bioinformatics analysis

Tumor RNA was extracted using RNeasy Mini Kit (cat no. 74106, Qiagen) for samples in tissuetech (n=98) and Allprep DNA/RNA FFPE (cat no. 80234, Qiagen) for FFPE samples (n=64). Library preparation was performed using KAPA HyperPrep kit (RiboErase HMR, Roche) followed by sequencing on an Illumina Novaseq6000. Salmon (17) was used to quantify the amount of each transcript using annotation from Gencode release 33 on genome assembly GRCh38 and transcript-level estimates were imported and summarized at gene-level using the R package tximport (v1.20). ComBat-seq was used to adjust for batch effects between fresh-frozen and FFPE samples using the R package sva (v3.42). Samples with less than five million mapped reads were excluded and genes not expressed in at least 25% of samples were removed, resulting in 131/162 tumors. Adjusted counts were normalized using edgeR (v3.34.1): effective library sizes were calculated using the trimmed mean of *M* values methods and counts were transformed to log2-counts-per-million. All tumors were classified according to the six consensus classes of MIBC (18) using the R package consensusMIBC (v1.1). Gene set enrichment analysis was performed using the R package fgsea (v1.20). Log-fold changes of genes between groups were estimated using genewise negative binomial generalized linear models (edgeR v3.36) and the Hallmark gene sets which were extracted from the Molecular Signatures Database using msigdbr (v7.5.1).

### Statistical analysis

Survival curves were compared using the Kaplan-Meier method. Hazard ratios (HR), associated 95% confidence intervals (CI) and *p*-values were calculated using Cox regression analysis (R packages survminer v0.4.9 and survival v3.2.13). Kruskal Wallis test, Wilcoxon rank sum test, Fisher’s exact test and Pearson’s Chi-squared test were used to determine statistically significant associations. ctDNA growth rates (slow rise vs fast rise) were calculated using a log-linear regression fitted to each patient based on ctDNA level as a function of time before clinical recurrence detection. The ctDNA growth rates were estimated from the slope of the regression lines. Analysis was performed using the R statistical environment (v4.1.2).

### Data availability

The raw sequencing data generated in this study are not publicly available as this compromises patient consent and ethics regulations in Denmark. Processed non-sensitive data are available upon reasonable request from the corresponding author.

## Results

### Cohort characteristics

We updated ctDNA data and clinical follow-up for 68 patients with MIBC who received NAC prior to RC (5). All patients were monitored longitudinally with plasma ctDNA testing. Additional ctDNA analysis of 56 plasma samples collected after RC was performed for 17 patients, resulting in a total of 712 plasma samples collected between 2014 and 2019 (**Supplementary Figure 1**). Patients had an updated median follow-up of 68 months after RC and an observed recurrence rate of 28% (18/65 patients).

In addition, a cohort of 102 patients with MIBC who did not receive NAC before RC were retrospectively selected from our biobank for ctDNA analysis. Plasma samples procured before RC (110 samples from 101 patients) and after RC (43 samples from 35 patients) between 2001 and 2016 were included (**Supplementary Figure 2**). Patients had a median follow-up of 72 months after RC and an observed recurrence rate of 44% (44/100 patients). Clinical characteristics differed between cohorts as patients in the NAC-treated, prospective cohort were collected consecutively while the patients in the NAC-naïve cohort were retrospectively selected to include a similar number of patients with and without metastatic disease within the cohort (**Table 1**).

**Table 1:** Clinical characteristics of patient cohorts

|  | NAC-treated cohort<br>(N=68)<sup>1</sup> | NAC-naïve cohort<br>(N=102)<sup>1</sup> | p-value<sup>2</sup> |
| --- | --- | --- | --- |
| Age, years, median (IQR) | 67 (59, 71) | 69 (62, 75) | 0.009 |
| Sex |  |  | 0.53 |
| Female | 12 (18%) | 22 (22%) |  |
| Male | 56 (82%) | 80 (78%) |  |
| T-stage, TURBT |  |  |  |
| Ta | 0 (0%) | 1 (1%) | 0.001 |
| T1 | 4 (5.9%) | 22 (22%) |  |
| T2 | 54 (79%) | 75 (74%) |  |
| T3 | 1 (1.5%) | 0 (0%) |  |
| T4 | 9 (13%) | 3 (3%) |  |
| N-stage, TURBT |  |  |  |
| N0 | 58 (85%) |  |  |
| N1 | 7 (10%) |  |  |
| N2 | 3 (4.4%) |  |  |
| T-stage, RC |  |  | <0.001 |
| T0/CIS/Ta | 44 (67%) | 27 (26%) |  |
| T1 | 3 (4.5%) | 8 (7.8%) |  |
| T2 | 6 (9.1%) | 23 (23%) |  |
| T3 | 9 (14%) | 35 (34%) |  |
| T4 | 4 (6.1%) | 9 (8.8%) |  |
| N-stage, RC |  |  | 0.059 |
| N0 | 59 (88%) | 79 (80%) |  |
| N1 | 2 (3%) | 13 (13%) |  |
| N2 | 3 (4.5%) | 6 (6.1%) |  |
| N3 | 3 (4.5%) | 1 (1%) |  |
| Pathological downstaging after NAC | 44 (66%) |  |  |
| Complete response after NAC | 42 (63%) |  |  |
| Recurrence | 18 (28%) | 44 (44%) | 0.035 |
| 12-month recurrence | 10 (15%) | 29 (30%) | 0.037 |
| RFS, months, median (IQR) | 24 (23, 25) | 24 (7, 32) | 0.10 |
| 2-year survival | 57 (86%) | 74 (73%) | 0.038 |
| 5-year survival | 48 (73%) | 58 (57%) | 0.049 |
| OS, months, median (IQR) | 68 (40, 81) | 72 (17, 92) | 0.042 |

<sup>1</sup>Median (IQR); n (%). <sup>2</sup>Wilcoxon rank sum test; Person's Chi-squared test; Fisher's exact test; Cox regression analysis. NAC=neoadjuvant chemotherapy; TURBT=transurethral resection of bladder tumor; CIS=carcinoma in situ; RC=radical cystectomy; RFS=recurrence-free survival; OS=overall survival.

### NAC-treated patient cohort: ctDNA detection is prognostic of outcomes

In this study with the updated clinical follow-up for the NAC-treated patients, ctDNA status remained highly prognostic of patient outcomes: at diagnosis before NAC (RFS: HR=15.6, 95%CI=3.5-69, *p*<0.0001; OS: HR=8.9, 95%CI=2.9-27.3, *p*=0.0001), after NAC prior to RC (RFS: HR=15.2, 95%CI=5-46.8, *p*<0.0001; OS: HR=9, 95%CI=3.6-22.6, *p*<0.0001), after RC (accumulated ctDNA status; RFS: HR=39.6, 95%CI=8.9-174.9, *p*<0.0001; OS: HR=15.4, 95%CI=5.8-40.8, *p*<0.0001) and within one year after RC (accumulated ctDNA status; RFS: HR not computable, log-rank p<0.0001; OS: HR=62.3, 95%CI=16-242.5, *p*<0.0001; **Figure 1A-L**). Notably, all patients with disease recurrence within the first year after RC were ctDNA-positive at diagnosis before NAC (**Figure 1C**). Five additional patients experienced metastatic relapse 20 to 61 months after RC with the extended follow-up. Of these, three patients (4422, 4479 and 4496) were ctDNA positive before NAC and during surveillance within the first two years after RC. Thereby, using full follow-up during surveillance after RC, we observe an overall recurrence rate of 100% (16/16) for ctDNA positive patients and 4.1% (2/49) for ctDNA negative patients (accumulated ctDNA status; 89% sensitivity, 100% specificity). For the remaining two patients (3889 and 3997) being ctDNA negative after RC but experiencing metastatic relapse, the last available plasma samples were collected 1,097 and 463 days before the clinical relapse, respectively. Restricting the analysis to include two years of follow-up after the last ctDNA analysis resulted in a recurrence rate of 94% (15/16) for ctDNA positive patients and 2% (1/49) for ctDNA negative patients (accumulated ctDNA status; 94% sensitivity, 98% specificity; **Figure 1I**).

**Figure 1.**
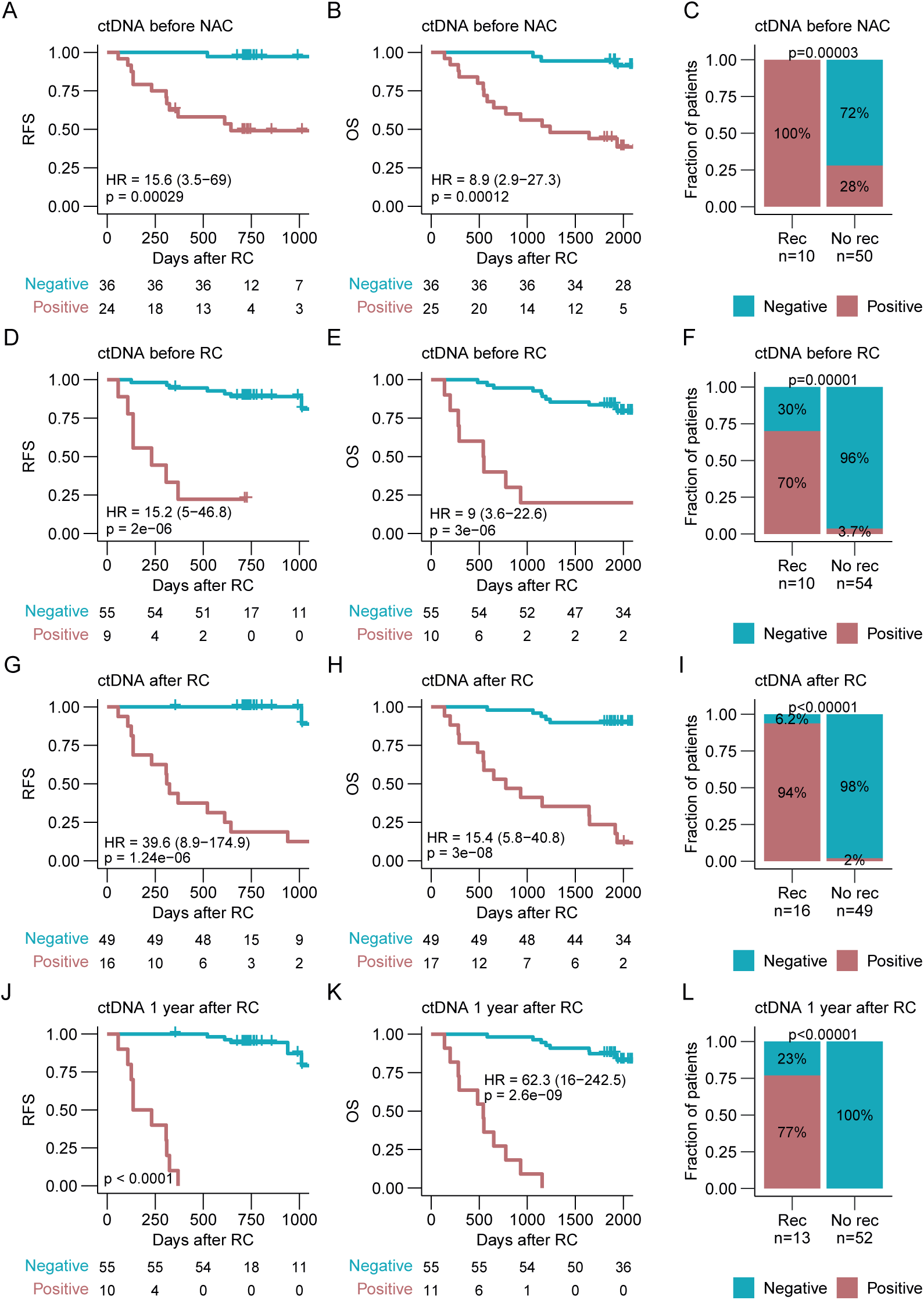
ctDNA detection for prognosis assessment in patients treated with NAC. **A,** Kaplan-Meier survival analysis of recurrence-free survival (RFS) and plasma ctDNA status before neoadjuvant chemotherapy (NAC). **B,** Kaplan-Meier survival analysis of overall survival (OS) and plasma ctDNA status before NAC. **C,** Association between plasma ctDNA status before NAC and recurrence status within one year after radical cystectomy (RC) including only patients with at least two years of follow-up after RC. **D,** Kaplan-Meier survival analysis of RFS and plasma ctDNA status before RC. **E,** Kaplan-Meier survival analysis of OS and plasma ctDNA status before RC. **F,** Association between plasma ctDNA status before RC and recurrence status within one year after RC including only patients with at least two years of follow-up after RC. **G,** Kaplan-Meier survival analysis of RFS and accumulated plasma ctDNA status after RC. **H,** Kaplan-Meier survival analysis of OS and accumulated plasma ctDNA status after RC. **I,** Association between accumulated plasma ctDNA status after RC and recurrence status within two years after the last plasma sample was analyzed for ctDNA. Only patients with at least two years of follow-up after RC were included. **J,** Kaplan-Meier survival analysis of RFS and accumulated plasma ctDNA status within one year after RC. **K,** Kaplan-Meier survival analysis of OS and accumulated plasma ctDNA status within one year after RC. **L,** Association between accumulated plasma ctDNA status within one year after RC and recurrence status within two years after the last plasma sample was analyzed including only patients with at least two years of follow-up after RC. Hazard ratios (HR) and associated 95% confidence intervals (CI) and p-values are displayed on each Kaplan-Meier plot (cox regression analysis). Significant statistical difference between ctDNA status and recurrence was determined using Fisher’s exact test.

For patients with metastatic relapse and detectable ctDNA (n=16), ctDNA analysis during surveillance after RC had a median lead-time of 123 days over radiographic imaging (range: −83 to 1478 days, *p*=0.005, full follow-up included) (**Figure 2A**). Importantly, ctDNA analysis had a median lead-time of 118 days over radiographic imaging (*p*=0.018) when restricting the analysis to patients with simultaneous plasma samples and imaging (imaging performed +/-1 month of the ctDNA analysis, n=13). We found an increase in ctDNA levels before clinical relapse for the 10 patients with metastatic relapse and ≥2 consecutive ctDNA positive plasma samples available before/after clinical relapse (**Figure 2B**). Interestingly, patients with longer lead-times (>200 days) had a slower rise in ctDNA levels (mean slope of 0.004) compared to patients with shorter lead-times (mean slope of 0.02; including patients with ≥2 consecutive ctDNA positive plasma samples within 365 days before and 30 days after their clinical relapse; **Figure 2C**).

**Figure 2.**
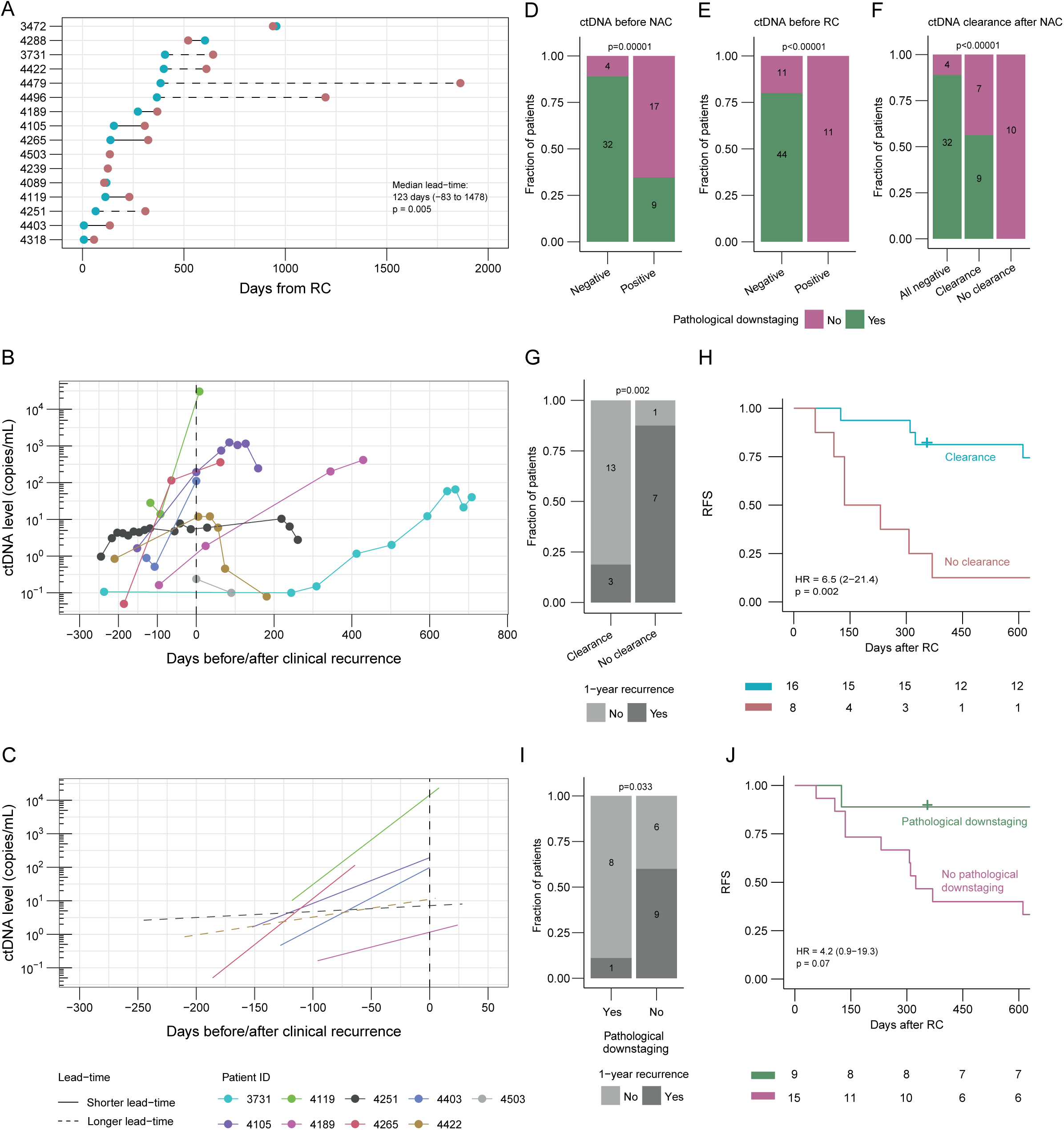
ctDNA measurements for monitoring relapse and treatment response. **A,** Lead time in days between molecular recurrence (ctDNA positivity) and clinical recurrence (radiographic imaging positive). Statistical significance was calculated using paired Wilcoxon rank sum test. Longer lead-time was defined as >200 days between molecular and clinical recurrence. **B,** ctDNA levels at the time of clinical recurrence (radiographic imaging positive, time point zero) for patients having at least two plasma samples analyzed for ctDNA at the time of their clinical relapse. **C,** Linear regression lines of ctDNA levels at the time of clinical recurrence (radiographic imaging positive, time point zero) for patients having at least two plasma samples analyzed for ctDNA at the time of their clinical relapse. Longer lead-time was defined as >200 days between molecular and clinical recurrence. **D,** Association between ctDNA status before neoadjuvant chemotherapy (NAC) and pathological downstaging. **E,** Association between ctDNA status before radical cystectomy (RC) and pathological downstaging. **F,** Association between ctDNA clearance after NAC and pathological downstaging. **G,** Association between ctDNA clearance after NAC and recurrence status within one year after RC for patients being ctDNA positive before NAC. **H,** Kaplan-Meier survival analysis of recurrence-free survival (RFS) and ctDNA clearance after NAC for patients being ctDNA positive before NAC. **I,** Association between pathological downstaging and recurrence status within one year after RC for patients being ctDNA positive before NAC. **J,** Kaplan-Meier survival analysis of RFS and pathological downstaging for patients being ctDNA positive before NAC. Hazard ratios (HR) and associated 95% confidence intervals (CI) and p-values are displayed on each Kaplan-Meier plot (cox regression analysis). Significant statistical difference between categorical variables was determined using Fisher’s exact test.

### ctDNA measurements for monitoring treatment response

As previously described, pathological downstaging to a non-invasive stage (≤pTa,CIS,N0) after NAC was observed for 66% of patients (44/67) (5). For patients without pathological downstaging, only 38% (8/21) had metastatic relapse within the first year after RC, indicating that pathological downstaging is suboptimal for evaluating treatment efficacy. Of the 36 patients who were ctDNA negative at diagnosis of MIBC, 89% (32/36) achieved pathological downstaging (**Figure 2D**). Likewise, 80% (44/55) of patients who were ctDNA negative after NAC prior to RC achieved pathological downstaging, whereas none of the ctDNA positive patients had pathological downstaging (**Figure 2E**). When evaluating ctDNA dynamics (measurements before and after NAC), 56% (9/16) of patients with ctDNA clearance achieved pathological downstaging while none of the patients persistently positive achieved pathological downstaging (**Figure 2F**). For patients who were ctDNA positive before treatment, clearance of ctDNA was significantly associated with disease relapse within one year after RC (*p*=0.002; **Figure 2G**) and full follow-up RFS (HR=6.5, 95%CI=2-21.4, *p*=0.002; **Figure 2H**). Pathological downstaging was significantly associated with disease relapse within one year (*p*=0.03; **Figure 2I**), but not RFS using the same patient subset (**Figure 2J**). Of note, multivariable Cox regression analysis revealed that both ctDNA positivity before RC (HR=5.4, 95%CI=1.5-19.1, *p*=0.009) and absence of ctDNA clearance after NAC (HR=4.7, 95%CI=1.2-18.8, *p*=0.029) were independently associated with worse RFS when adjusted for pathological downstaging (**Supplementary Table 1**), indicating that ctDNA analysis might be an appropriate tool to evaluate treatment efficacy as well as to identify high risk patients before RC is performed.

### NAC-naïve cohort: ctDNA detection is prognostic of outcomes

For the NAC-naïve patients, the presence of ctDNA was a strong prognostic predictor of patient outcomes: at diagnosis before RC (RFS: HR=3.4, 95%CI=1.7-6.8, *p*=0.0005) and after RC (accumulated ctDNA status; RFS: HR=17.8, 95%CI=3.9-81.2, *p*=0.0002; **Figure 3A-F**). In this cohort, accumulated ctDNA status during disease surveillance after RC identified metastatic relapse with 82% sensitivity and 94% specificity (**Figure 3F**; recurrence evaluation within two years after the last plasma sample was analyzed for ctDNA). We hypothesized that patients without detectable ctDNA at diagnosis do not need NAC and would therefore have a similar recurrence rate as ctDNA negative, NAC-treated patients. However, when comparing recurrence rates within one year after RC for patients who were ctDNA negative at diagnosis between the two cohorts, we observed a recurrence rate of 10% (5/50) in NAC-naïve patients compared to 0% in NAC-treated patients. Notably, the difference in 1-year recurrence rates (*p*=0.07) or RFS (*p*=0.05) was not statistically significant (**Figure 3G**). The RFS of ctDNA positive patients in the NAC-naïve cohort was not shorter compared to ctDNA positive patients in the NAC-treated cohort and we observed no difference in 1-year recurrence rates (**Figure 3H**).

**Figure 3.**
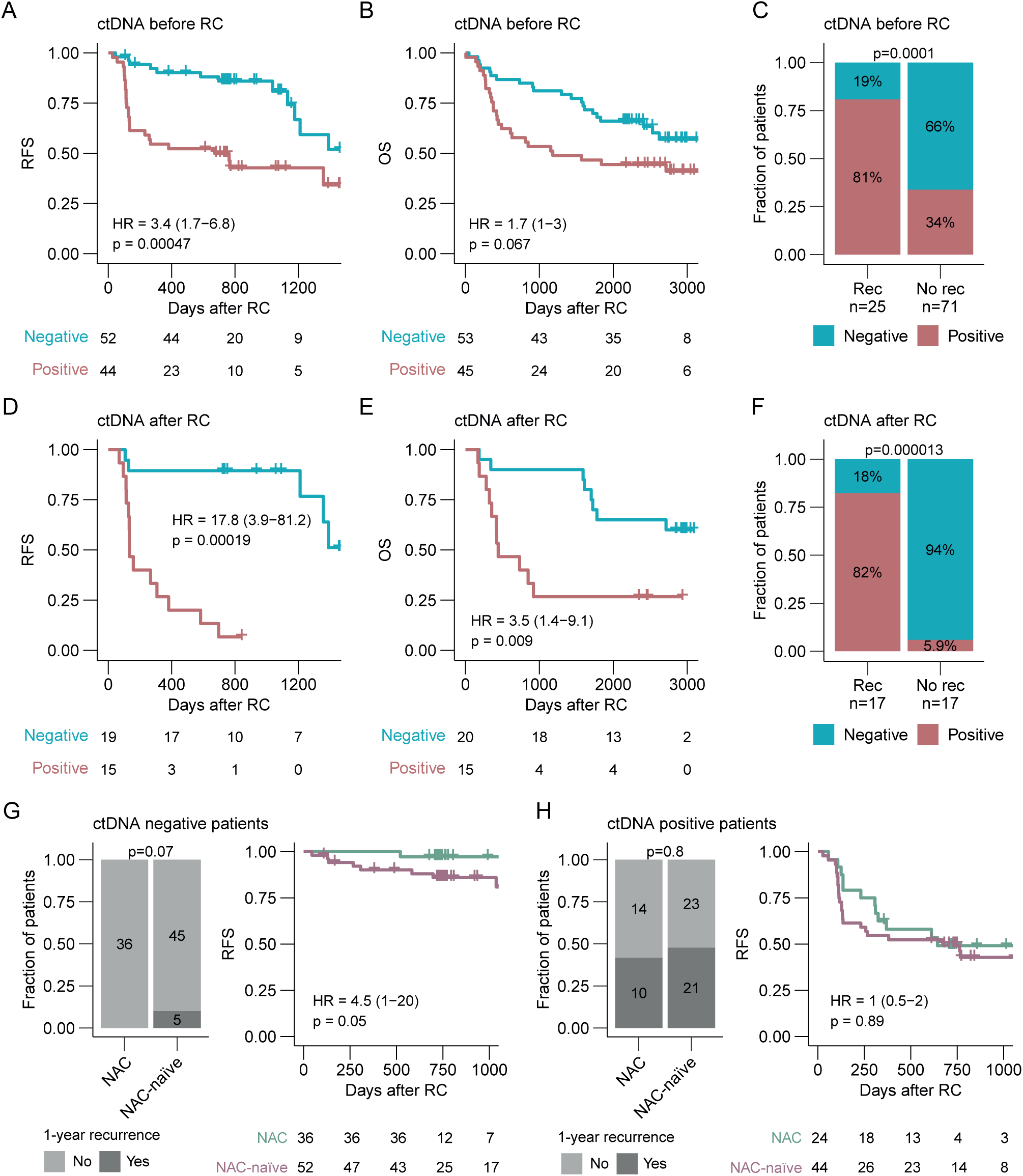
ctDNA detection for prognosis assessment in patients not treated with NAC. **A,** Kaplan-Meier survival analysis of recurrence-free survival (RFS) and plasma ctDNA status before radical cystectomy (RC). **B,** Kaplan-Meier survival analysis of overall survival (OS) and plasma ctDNA status before RC. **C,** Association between plasma ctDNA status before RC and recurrence status within one year after RC including only patients with at least two years of follow-up after RC. **D,** Kaplan-Meier survival analysis of RFS and plasma ctDNA status after RC. **E,** Kaplan-Meier survival analysis of OS and accumulated plasma ctDNA status after RC. **F,** Association between plasma ctDNA status after RC and recurrence status within two years after the last plasma sample was analyzed for ctDNA. Only patients with at least two years of follow-up after RC were included. **G,** 12-month recurrence rates (left) and Kaplan-Meier survival analysis of RFS (right) for baseline ctDNA negative patients in the NAC-treated and NAC-naïve cohorts. **H,** 12-month recurrence rates (left) and Kaplan-Meier survival analysis of RFS (right) for baseline ctDNA positive patients in the NAC-treated and NAC-naïve cohorts. Hazard ratios (HR) and associated 95% confidence intervals (CI) and p-values are displayed on each Kaplan-Meier plot (cox regression analysis). Significant statistical difference between ctDNA status and recurrence was determined using Fisher’s exact test.

Due to the collection procedure of plasma samples in the NAC-naïve cohort, the volume of plasma obtained, the amount of extracted cfDNA and the library input were significantly lower compared to the NAC-treated cohort, thus potentially reducing overall sensitivity of detecting ctDNA in the NAC-naïve cohort (**Supplementary Figure 3A-C**). Five patients in the NAC-naïve cohort did not have detectable ctDNA at the pre-RC time point despite having metastatic relapse within 1 year after RC, but the quality of the samples from these patients was not found to be significantly lower compared to the remaining samples in the cohort (**Supplementary Figure 3D-F**). However, despite the overall lower quality of samples in the NAC-naïve cohort, a strong prognostic value of ctDNA testing remained (**Figure 3**).

### Clinical and biological characterization of ctDNA shedding tumors

To study features of baseline ctDNA positivity, we combined all analyzed tumors and utilized the pre-NAC ctDNA status for NAC-treated patients and the pre-RC ctDNA status for NAC-naïve patients. In agreement with previous findings, ctDNA detection at baseline was associated with higher tumor stage (*p*=0.008) and tumor size (*p*=0.016) at TURBT (**Figure 4A-B**). Smoking status of the patients was not associated with ctDNA positivity at baseline (**Figure 4C**). Using the WES data from tumors, *TP53* was the only driver gene showing a higher mutation rate in ctDNA shedding tumors (*p*=0.007; **Supplementary Figure 4A**). We found no association between the number of somatic variants or contribution of mutational signatures and baseline ctDNA positivity (**Supplementary Figure 4B-D**).

**Figure 4.**
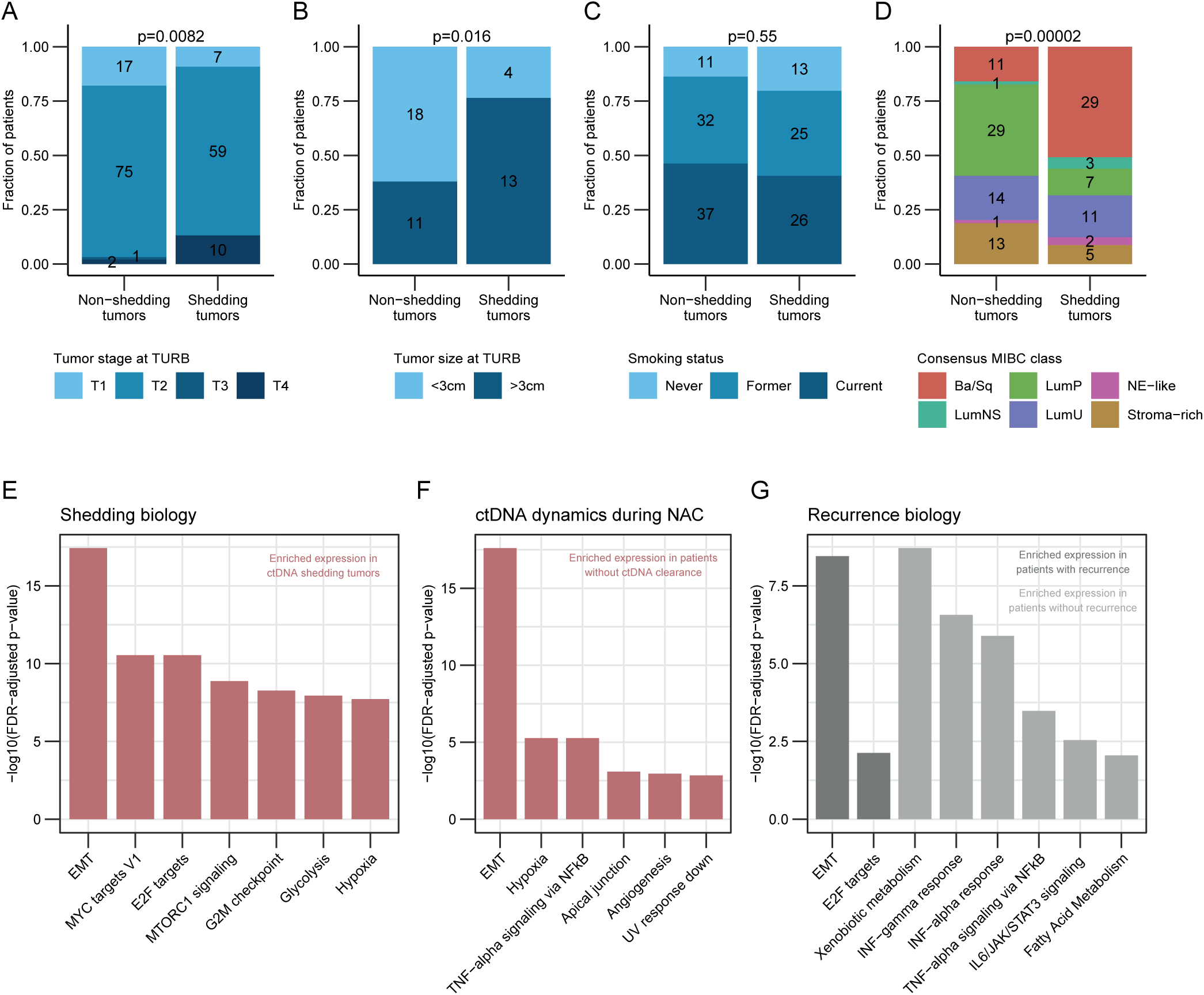
Biological characterization of ctDNA shedding tumors. **A,** Association between baseline ctDNA status and tumor stage at TURBT including both patient cohorts. **B,** Association between baseline ctDNA status and tumor size at TURBT including both patient cohorts. **C,** Association between baseline ctDNA status and smoking status of patients including both cohorts. **D,** Association between baseline ctDNA status and consensus classification of tumors including both patient cohorts. **E,** Gene set enrichment analysis of tumors using the Hallmark pathways comparing baseline ctDNA shedding and non-shedding patients from both cohorts. Only gene sets with a normalized enrichment score >2 are shown. **F,** Gene set enrichment analysis of tumors using the Hallmark pathways comparing patients with and without ctDNA clearance after neoadjuvant chemotherapy. **G,** Gene set enrichment analysis of tumors using the Hallmark pathways comparing patients with and without recurrence within one year after radical cystectomy. Statistical significance of ctDNA status and other categorical variables was calculated using Fisher’s exact test. Gene sets in E-G were ordered by decreasing q-values.

To explore ctDNA shedding biology further, total RNA-sequencing was performed on 162 tumors of which 131 passed quality control filtering. Tumors were classified according to the six MIBC consensus classes (18) and more Basal/Squamous (Ba/Sq) tumors were found among the ctDNA positive patients (*p*<0.0001; **Figure 4D**), possibly reflecting the higher aggressiveness of Ba/Sq tumors compared to the luminal subtypes. For ctDNA shedding tumors, ctDNA levels were not associated with tumor subtype classifications (**Supplementary Figure 4E**). In both cohorts, gene set enrichment analysis using the Hallmark gene sets revealed an enrichment of oncogenic pathways, namely epithelial-to-mesenchymal transition (EMT; *q*<0.0001), hypoxia (*q*<0.0001) and cell-cycle associated gene sets (E2F targets, *q*<0.0001 and G2M checkpoint, *q*<0.0001) in tumors from ctDNA positive patients (**Figure 4E**; **Supplementary Figure 4F**). This may reflect a more aggressive cancer phenotype with metastatic abilities of ctDNA shedding tumors. Gene sets upregulated in patients who did not have ctDNA clearance after NAC (n=7 patients) compared to patients with ctDNA clearance (n=11 patients) included EMT (*q*<0.0001), hypoxia (*q*<0.0001) and TNFα signaling (*q*<0.0001; **Figure 4F**). When using clinical relapse within one year after RC as endpoint instead of ctDNA status, we found enrichment of the EMT and E2F targets in patients with metastatic relapse (n=32 patients) whereas enrichment of anti-tumor immune pathways, including IFNα (*q*<0.0001) and IFNγ response (*q*<0.0001) was observed among patients without relapse (n=91; **Figure 4G**).

## Discussion

Assessment of ctDNA is a promising minimally-invasive blood-based biomarker in bladder cancer and here we underline its prognostic value in NAC-treated patients with >5 years of follow-up after RC. Accumulated ctDNA analysis after RC identified metastatic relapse with a sensitivity of 94% and specificity of 98%, highlighting the potential of ctDNA-guided prognostication and patient management using a personalized, tumor-informed assay. We observed a positive lead-time of ctDNA-based relapse detection of 123 days over radiographic imaging, and importantly we still observed a positive lead-time of 118 days when restricting the analysis to patients with simultaneous ctDNA analysis and radiographic imaging. The association between longer lead-times and a slower rise in ctDNA levels reflects more indolent underlying tumor biology and supports the need for a frequent, quantitative assessment in this setting compared to a binary evaluation of ctDNA presence.

We included analysis of a NAC-naïve patient cohort to evaluate the prognostic value of ctDNA in this setting. We found that the presence of ctDNA was highly associated with worse RFS as expected. When comparing outcomes across the patient cohorts, we observed a recurrence rate within one year after RC of 0% and 10% (non-significant) among baseline ctDNA negative patients in the NAC-treated and NAC-naïve cohort, respectively. Although the difference in RFS was not statistically significant, we hypothesize that the higher recurrence rate of the ctDNA negative patients in the NAC-naïve cohort could be caused by the presence of non-detected micrometastases or dormant carcinoma cells not eradicated by NAC. Of note, patients in the NAC-naïve cohort were selected to represent an equal number of patients with and without metastatic disease within the cohort, explaining the overall higher number of recurrence events observed in this cohort compared to the prospectively collected cohort of NAC-treated patients. Furthermore, the plasma sample quality and volume in the NAC-naïve cohort were lower compared to the samples in the NAC-treated cohort, potentially impacting sensitivity. We speculate that this might explain ctDNA negativity at baseline for the five patients experiencing metastatic relapse within 12 months after RC. Finally, the higher recurrence rate of the ctDNA negative patients in the NAC-naïve cohort might also be caused by the presence of non-detected micrometastases or dormant carcinoma cells not eradicated by NAC. A future randomized clinical trial is needed to establish whether baseline ctDNA negative patients could potentially avoid NAC and its associated toxicity, or whether NAC is still indicated despite no ctDNA detection. No difference in RFS of ctDNA positive patients was observed between patient cohorts; however, due to discrepancies between the cohorts, the interpretation of a cross-cohort comparison in both ctDNA negative and positive patients has its limitations. There is potential to explore whether escalation of treatment will improve the outcome of the high-risk patients who are ctDNA positive at diagnosis and do not respond to NAC (remain ctDNA positive after NAC or become ctDNA positive shortly after RC).

Assessment of ctDNA as treatment response parameter has previously been explored in MIBC (5,6). Here we found that ctDNA status and ctDNA dynamics during NAC were highly associated with pathological downstaging. Furthermore, ctDNA status before RC and ctDNA dynamics during NAC both outperformed pathological downstaging in predicting treatment efficacy and patient outcomes after RC. Defects in DNA damage repair (DDR) genes have been shown to increase sensitivity to chemotherapy in bladder cancer (19–22) and several ongoing clinical trials are investigating bladder sparing for patients with alterations in DDR genes and complete pathological response following NAC (23–25). The combined approach of ctDNA testing and assessment of DDR pathway alterations could potentially provide a refined selection of patients for bladder preserving protocols. However, other studies have observed no predictive power of DDR gene mutations (5,26). A recent publication, where urinary tumor-derived DNA was evaluated for 56 out of the 68 patients in the NAC-treated cohort (27), highlighted that a combined analysis of urine- and plasma samples in pre-RC setting may provide further strength to identify low risk patients potentially eligible for bladder sparing approaches.

We sought to characterize the underlying biology of ctDNA shedding tumors and generally observed a more aggressive phenotype with enrichment of EMT and cell-cycle associated gene sets. These findings are in line with previous observations in lung cancer where ctDNA positive adenocarcinomas also showed upregulation of proliferation and cell-cycle associated gene sets (13). In addition, we found more tumors of the Ba/Sq subtype among patients being ctDNA positive at baseline. This has previously been observed in MIBC (28) and a previous study on lung cancer found a higher ctDNA detection rate for squamous tumors compared to adenocarcinomas (11,13), which was suggested to be caused by the more necrotic profile of squamous tumors. In summary, these results suggest that presence of EMT in the primary tumor might influence ctDNA shedding and that a high proliferation rate and overall necrotic profile increases the continuous shedding of ctDNA into circulation, making ctDNA detection in these patients more likely. When using ctDNA dynamics to evaluate treatment response, we found enrichment of the hypoxia gene set amongst others in tumors from patients without ctDNA clearance during NAC. Hypoxia has been suggested to enhance chemoresistance and the metastatic potential of cancer cells (29), further linking ctDNA dynamics during treatment and lack of response to therapy.

Implementation of ctDNA analysis in the management of patients with MIBC could potentially solve several clinical challenges, including early prognostication of patients, monitoring of treatment response, early detection of metastatic relapse and potentially selection of patients for bladder preserving protocols. Clinical trials evaluating ctDNA-guided adjuvant treatment with atezolizumab are currently ongoing (30,31), but additional randomized trials evaluating ctDNA-stratified therapeutic and bladder preserving approaches are needed to further elucidate the full clinical potential of ctDNA assessment.

## Acknowledgements

We would like to thank all technical personnel at the Departments of Molecular Medicine, Urology and Oncology at Aarhus University Hospital, Denmark for sample handling and processing.

## References

1. Sung H, Ferlay J, Siegel RL, Laversanne M, Soerjomataram I, Jemal A, et al. Global Cancer Statistics 2020: GLOBOCAN Estimates of Incidence and Mortality Worldwide for 36 Cancers in 185 Countries. CA Cancer J Clin. 2021;71:209–49.

2. Witjes JA, Bruins HM, Cathomas R, Compérat EM, Cowan NC, Gakis G, et al. European Association of Urology Guidelines on Muscle-invasive and Metastatic Bladder Cancer: Summary of the 2020 Guidelines. Eur Urol. 2021;79:82–104.

3. Niedersüss-Beke D, Puntus T, Kunit T, Grünberger B, Lamche M, Loidl W, et al. Neoadjuvant Chemotherapy with Gemcitabine plus Cisplatin in Patients with Locally Advanced Bladder Cancer. Oncology. 2017;93:36–42.

4. Zargar H, Espiritu PN, Fairey AS, Mertens LS, Dinney CP, Mir MC, et al. Multicenter assessment of neoadjuvant chemotherapy for muscle-invasive bladder cancer. Eur Urol. 2015;67:241–9.

5. Christensen E, Birkenkamp-Demtröder K, Sethi H, Shchegrova S, Salari R, Nordentoft I, et al. Early Detection of Metastatic Relapse and Monitoring of Therapeutic Efficacy by Ultra-Deep Sequencing of Plasma Cell-Free DNA in Patients With Urothelial Bladder Carcinoma. J Clin Oncol. 2019;37:1547–57.

6. Powles T, Assaf ZJ, Davarpanah N, Banchereau R, Szabados BE, Yuen KC, et al. ctDNA guiding adjuvant immunotherapy in urothelial carcinoma. Nature. 2021;595:432– 7.

7. van Dorp J, Pipinikas C, Suelmann BBM, Mehra N, van Dijk N, Marsico G, et al. High- or low-dose preoperative ipilimumab plus nivolumab in stage III urothelial cancer: the phase 1B NABUCCO trial. Nat Med [Internet]. 2023; Available from: http://dx.doi.org/10.1038/s41591-022-02199-y

8. Bettegowda C, Sausen M, Leary RJ, Kinde I, Wang Y, Agrawal N, et al. Detection of circulating tumor DNA in early- and late-stage human malignancies. Sci Transl Med. 2014;6:224ra24.

9. Abbosh C, Birkbak NJ, Wilson GA, Jamal-Hanjani M, Constantin T, Salari R, et al. Phylogenetic ctDNA analysis depicts early-stage lung cancer evolution. Nature. 2017;545:446–51.

10. Abbosh C, Birkbak NJ, Swanton C. Early stage NSCLC - challenges to implementing ctDNA-based screening and MRD detection. Nat Rev Clin Oncol. 2018;15:577–86.

11. Sánchez-Herrero E, Serna-Blasco R, Robado de Lope L, González-Rumayor V, Romero A, Provencio M. Circulating Tumor DNA as a Cancer Biomarker: An Overview of Biological Features and Factors That may Impact on ctDNA Analysis. Front Oncol. 2022;12:943253.

12. Hsiehchen D, Espinoza M, Gerber DE, Beg MS. Clinical and biological determinants of circulating tumor DNA detection and prognostication using a next-generation sequencing panel assay. Cancer Biol Ther. 2021;22:455–64.

13. Abbosh C, Frankell AM, Harrison T, Kisistok J, Garnett A, Johnson L, et al. Tracking early lung cancer metastatic dissemination in TRACERx using ctDNA. Nature. 2023;616:553–62.

14. Herberts C, Annala M, Sipola J, Ng SWS, Chen XE, Nurminen A, et al. Deep whole-genome ctDNA chronology of treatment-resistant prostate cancer. Nature. 2022;608:199–208.

15. Harris PA, Taylor R, Thielke R, Payne J, Gonzalez N, Conde JG. Research electronic data capture (REDCap)--a metadata-driven methodology and workflow process for providing translational research informatics support. J Biomed Inform. 2009;42:377–81.

16. Harris PA, Taylor R, Minor BL, Elliott V, Fernandez M, O’Neal L, et al. The REDCap consortium: Building an international community of software platform partners. J Biomed Inform. 2019;95:103208.

17. Patro R, Duggal G, Love MI, Irizarry RA, Kingsford C. Salmon provides fast and bias-aware quantification of transcript expression. Nat Methods. 2017;14:417–9.

18. Kamoun A, de Reyniès A, Allory Y, Sjödahl G, Robertson AG, Seiler R, et al. A Consensus Molecular Classification of Muscle-invasive Bladder Cancer. Eur Urol. 2020;77:420–33.

19. Van Allen EM, Mouw KW, Kim P, Iyer G, Wagle N, Al-Ahmadie H, et al. Somatic ERCC2 mutations correlate with cisplatin sensitivity in muscle-invasive urothelial carcinoma. Cancer Discov. 2014;4:1140–53.

20. Plimack ER, Dunbrack RL, Brennan TA, Andrake MD, Zhou Y, Serebriiskii IG, et al. Defects in DNA Repair Genes Predict Response to Neoadjuvant Cisplatin-based Chemotherapy in Muscle-invasive Bladder Cancer. Eur Urol. 2015;68:959–67.

21. Teo MY, Bambury RM, Zabor EC, Jordan E, Al-Ahmadie H, Boyd ME, et al. DNA Damage Response and Repair Gene Alterations Are Associated with Improved Survival in Patients with Platinum-Treated Advanced Urothelial Carcinoma. Clin Cancer Res. 2017;23:3610–8.

22. Iyer G, Balar AV, Milowsky MI, Bochner BH, Dalbagni G, Donat SM, et al. Multicenter Prospective Phase II Trial of Neoadjuvant Dose-Dense Gemcitabine Plus Cisplatin in Patients With Muscle-Invasive Bladder Cancer. J Clin Oncol. 2018;36:1949–56.

23. Risk Enabled Therapy After Initiating Neoadjuvant Chemotherapy for Bladder Cancer (RETAIN) [Internet]. [cited 2023 May 16]. Available from: https://clinicaltrials.gov/ct2/show/NCT02710734

24. Gemcitabine, Cisplatin, Plus Nivolumab in Patients With Muscle-invasive Bladder Cancer With Selective Bladder Sparing [Internet]. [cited 2023 May 16]. Available from: https://clinicaltrials.gov/ct2/show/NCT03558087

25. Gemcitabine and Cisplatin Without Cystectomy for Patients With Muscle Invasive Bladder Urothelial Cancer and Select Genetic Alterations [Internet]. [cited 2023 May 16]. Available from: https://clinicaltrials.gov/ct2/show/NCT03609216

26. Taber A, Christensen E, Lamy P, Nordentoft I, Prip F, Lindskrog SV, et al. Molecular correlates of cisplatin-based chemotherapy response in muscle invasive bladder cancer by integrated multi-omics analysis. Nat Commun. 2020;11:4858.

27. Christensen E, Nordentoft I, Birkenkamp-Demtröder K, Elbæk SK, Lindskrog SV, Taber A, et al. Cell-Free Urine and Plasma DNA Mutational Analysis Predicts Neoadjuvant Chemotherapy Response and Outcome in Patients with Muscle-Invasive Bladder Cancer. Clin Cancer Res. 2023;29:1582–91.

28. Szabados B, Kockx M, Assaf ZJ, van Dam P-J, Rodriguez-Vida A, Duran I, et al. Final Results of Neoadjuvant Atezolizumab in Cisplatin-ineligible Patients with Muscle-invasive Urothelial Cancer of the Bladder. Eur Urol [Internet]. 2022; Available from: http://dx.doi.org/10.1016/j.eururo.2022.04.013

29. Wilson WR, Hay MP. Targeting hypoxia in cancer therapy. Nat Rev Cancer. 2011;11:393–410.

30. A Study of Atezolizumab Versus Placebo as Adjuvant Therapy in Patients With High-Risk Muscle-Invasive Bladder Cancer Who Are ctDNA Positive Following Cystectomy - Full Text View - ClinicalTrials.gov [Internet]. [cited 2023 May 16]. Available from: https://clinicaltrials.gov/ct2/show/NCT04660344

31. Treatment Of Metastatic Bladder Cancer at the Time Of Biochemical reLApse Following Radical Cystectomy - Full Text View - ClinicalTrials.gov [Internet]. [cited 2023 May 16]. Available from: https://clinicaltrials.gov/ct2/show/NCT04138628

