## Supplementary Figures and Tables for "Circulating tumor DNA analysis in advanced urothelial carcinoma: insights from biological analysis and extended clinical follow-up"

### Supplementary Figure 1

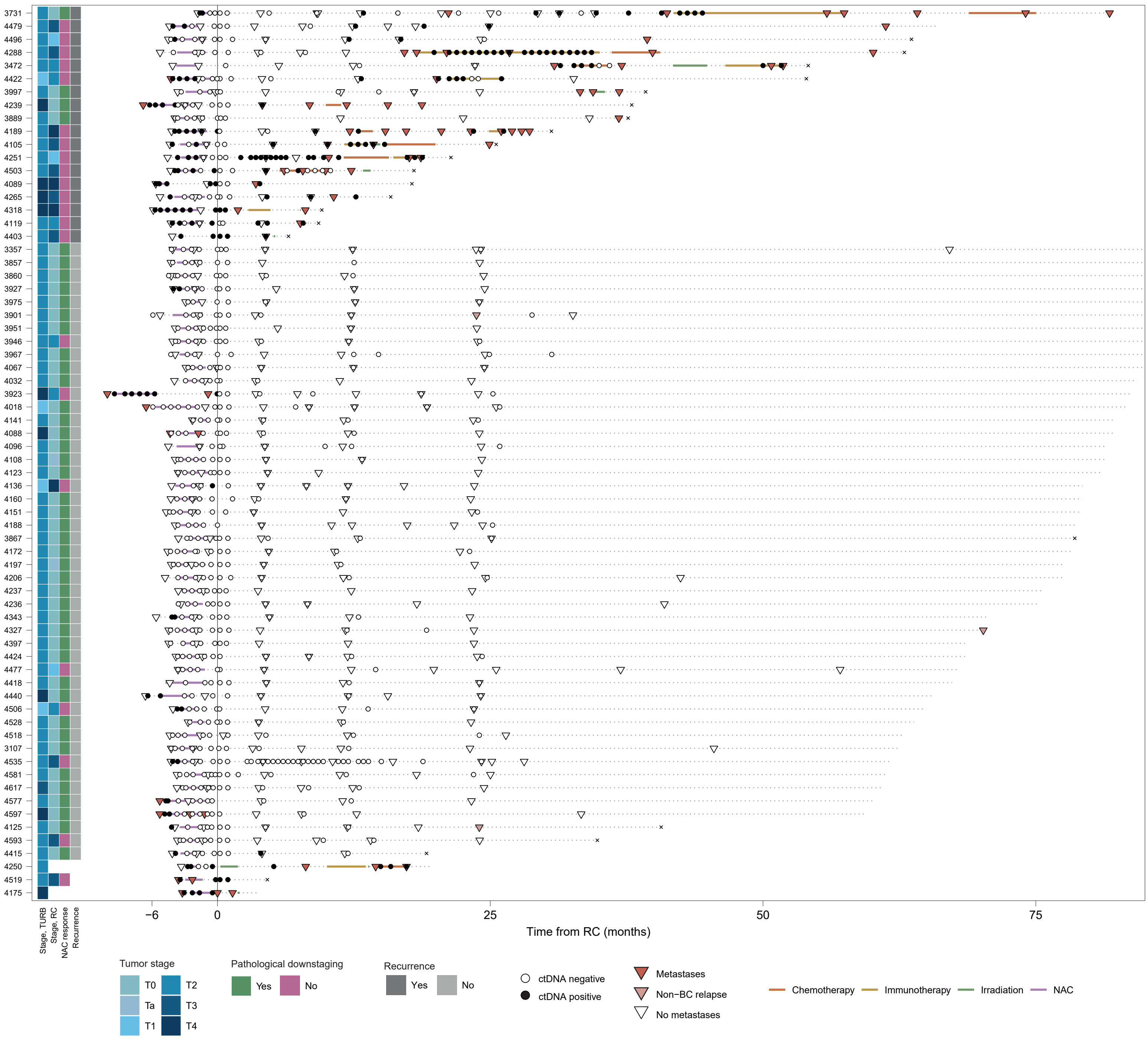

Longitudinal representation of circulating tumor DNA (ctDNA) results for analyzed samples in the NAC-treated patient cohort (n=68). Patients are ordered by decreasing overall survival within patients with and without recurrence.

### Supplementary Figure 2

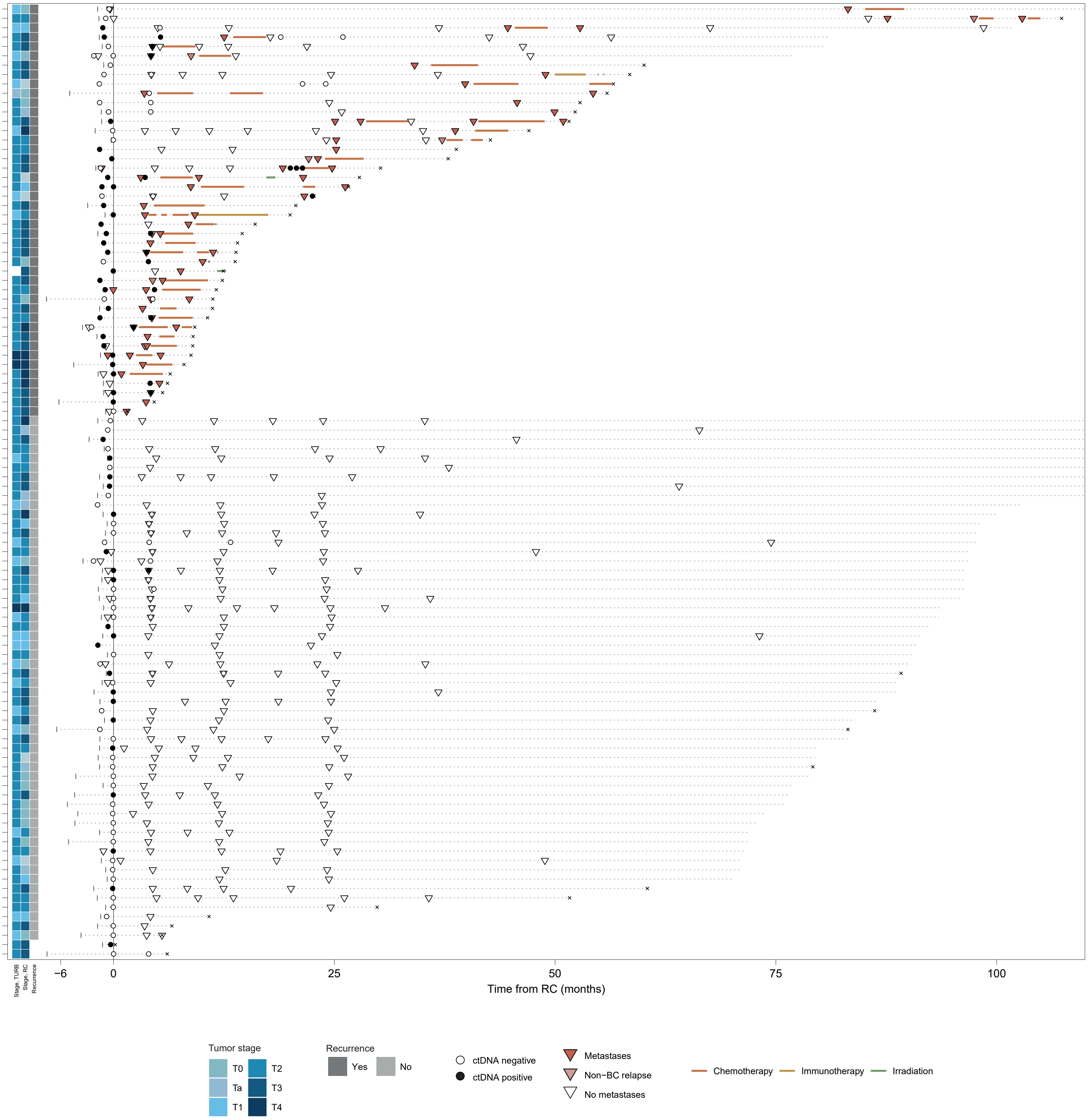

Longitudinal representation of circulating tumor DNA (ctDNA) results for analyzed samples in the NAC-naïve patient cohort (n=102). Patients are ordered by decreasing overall survival within patients with and without recurrence.

#### Supplementary Figure 3

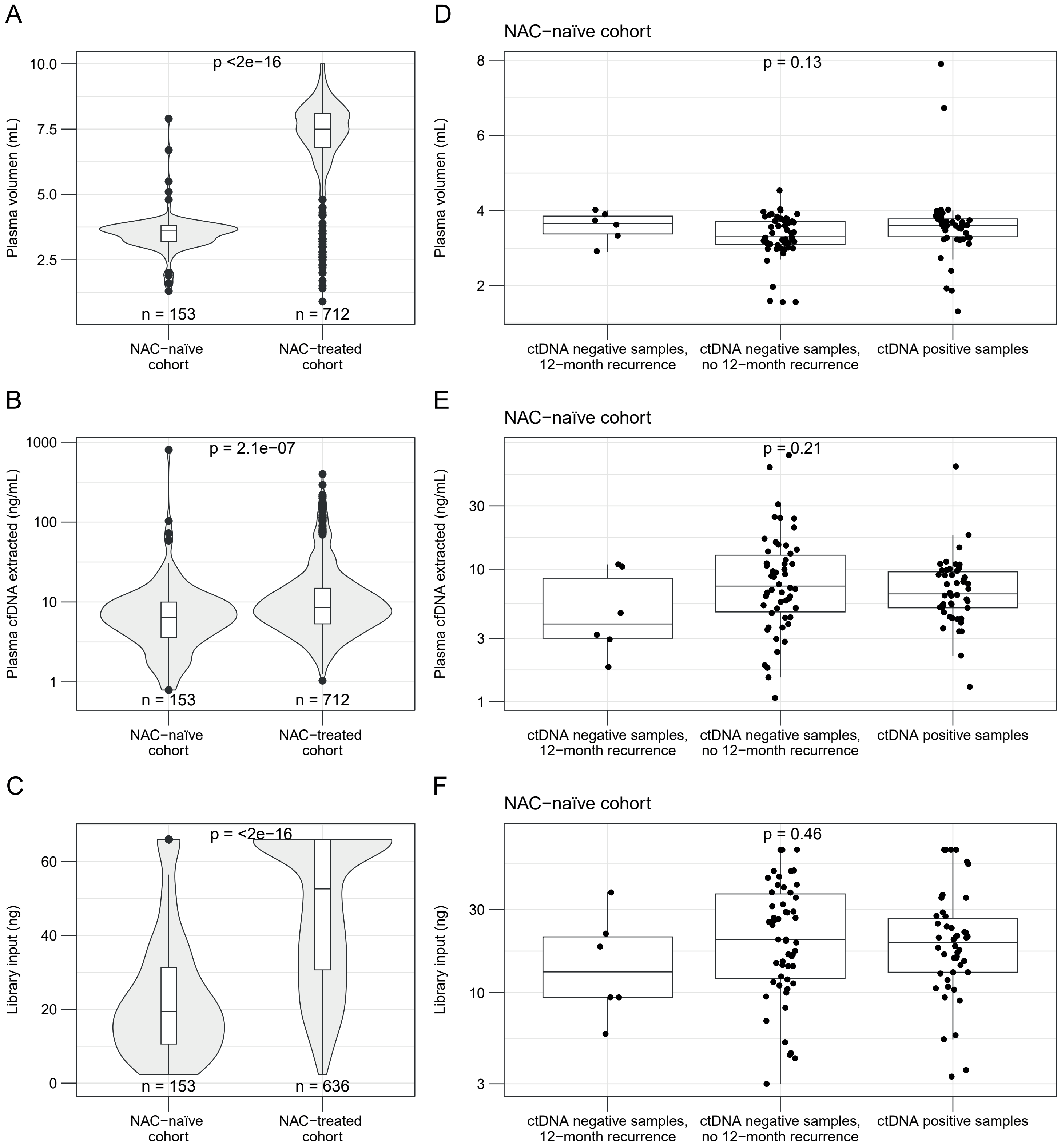

Quality metrics of plasma samples. **A,** Plasma volume of analyzed samples in the NAC-treated and NAC-naïve cohorts. **B,** Amount of plasma cell-free DNA (cfDNA) extracted from samples in the NAC-treated and NAC-naïve cohorts. **C,** Library input for the NAC-treated and NAC-naïve cohorts. **D,** Plasma volume of analyzed samples collected before radical cystectomy (RC) in the NAC-naïve cohort separated by ctDNA status and whether the patient had metastatic relapse within 12 months after RC for ctDNA negative samples. **E,** Amount of cfDNA extracted from samples collected before RC in the NAC-naïve cohort separated by ctDNA status and whether the patient had metastatic relapse within 12 months after RC for ctDNA negative samples. **F,** Library input for samples collected before RC in the NAC-naïve cohort separated by ctDNA status and whether the patient had metastatic relapse within 12 months after RC for ctDNA negative samples.

### Supplementary Figure 4

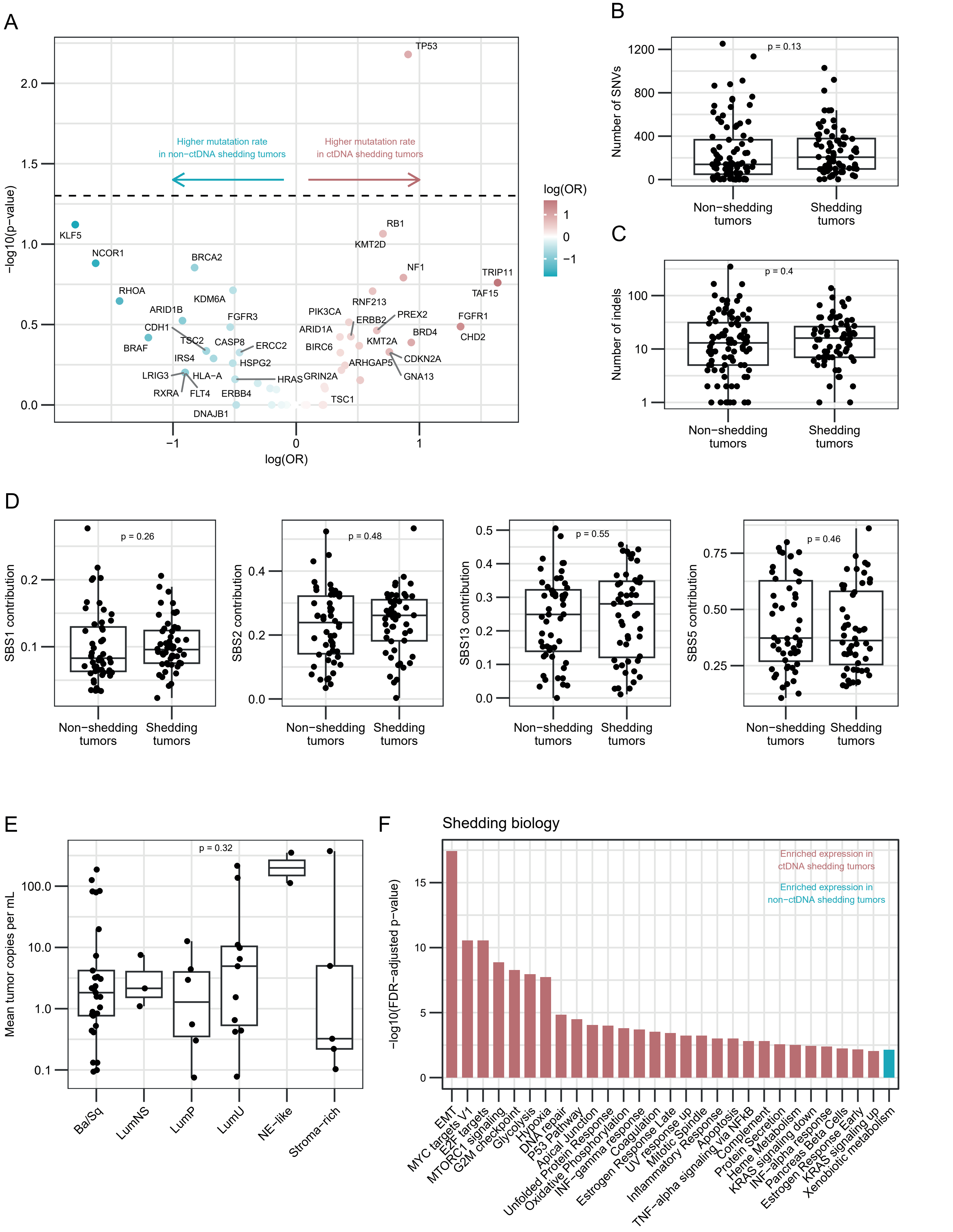

Biological characteristics of ctDNA shedding tumors. **A,** Volcano plot showing mutations associated with either ctDNA shedding or non-shedding tumors. Dashed horizontal line indicates p-values of 0.05 (non-adjusted). Statistical significance was assessed by Fisher's exact test. **B,** Association between single-nucleotide variations (SNVs) and ctDNA status at baseline including both cohorts. **C,** Association between indels and ctDNA status at baseline including both cohorts. **D,** Association between bladder cancer related mutational signatures and ctDNA status at baseline including both cohorts. **E,** Association between mean tumor copies per mL and consensus MIBC classification of primary tumor. **F,** Gene set enrichment analysis of tumors using the Hallmark pathways comparing baseline ctDNA shedding and non-shedding patients from both cohorts.

Supplementary Table 1: Univariate- and multivariable cox regression analysis

| **Recurrence-free survival** | | |
|---|---|---|
| | HR (95% CI) | *p*-value |
| **Univariate analysis** | | |
| Pathological downstaging (no vs yes; n=65, 18 events) | 9.2 (3.0-28.0) | 0.0001 |
| ctDNA status before RC (positive vs negative; n=64, 17 events) | 15.2 (4.9-46.8) | <0.0001 |
| ctDNA clearance during NAC (no vs yes; n=24, 14 events) | 6.5 (1.9-21.4) | 0.002 |
| **Multivariable model 1 (n=64, 17 events)** | | |
| Pathological downstaging (no vs yes) | 4.9 (1.4-17.9) | 0.015 |
| ctDNA status before RC (positive vs negative) | 5.4 (1.5-19.1) | 0.009 |
| **Multivariable model 2 (n=24, 14 events)** | | |
| Pathological downstaging (no vs yes) | 2.0 (0.3-11.8) | 0.46 |
| ctDNA clearance during NAC (no vs yes) | 4.7 (1.2-18.8) | 0.029 |

HR = hazard ratio; CI = confidence interval; NAC = neoadjuvant chemotherapy; RC = radical cystectomy.
